# External Validation of a Mathematical Model of Brain Health

**DOI:** 10.64898/2026.09.01.26361929

**Authors:** Halima Sadia, Nicolas Doyon, Simon Duchesne, Alzheimer’s Disease Neuroimaging Initiative

## Abstract

**Background:** Understanding the mechanisms underlying brain aging and age-related pathological changes is essential for advancing brain health research. Our group previously developed a mechanistic mathematical model of healthy brain Chamberland et al. (2024) that integrates key biological processes involved in normal aging, from which Alzheimer’s disease (AD)-related changes may emerge naturally.

**Objectives:** To characterize and validate this brain model by evaluating its sensitivity, calibrating its parameters, and assessing generalizability in independent populations.

**Methods:** The model represents the evolution of key biological processes associated with brain aging, including amyloid beta (A*β*), tau pathologies, neuroinflammation, and neuronal death. After identifying the 30 most influential parameters, we calibrated the model using cognitively normal (CN) participants from the AD Neuroimaging Initiative (ADNI) database (*n* = 211) by minimizing a loss function composed of three outcomes (A*β* plaques, tau tangles, and neuronal density). The calibrated model was then applied to the UK Biobank cohort (*n* = 35, 899) of normal controls (aged 44–82 years). The effects of sex and *APOE* were evaluated using stratified simulations.

**Results:** Parameter calibration significantly reduced the prediction errors for A*β* and tau. Neuronal density predictions showed strong agreement in the UK Biobank cohort. The variance decomposition identified *APOE* status as a major contributor to variability in A*β*.

**Conclusion:** Our validated brain health model links mechanistic pathways with population data and reproduces neuronal density patterns in an independent cohort. These findings support its use as a framework for studying brain aging and investigating how Alzheimer’s disease-related pathological changes may emerge with aging.

## Introduction

Alzheimer’s disease (AD) has become a major public health problem, with its burden expected to jeopardize health care systems as its prevalence is projected to increase due to an aging global population ^1^. However, understanding how the healthy brain changes with age is essential to characterize the biological mechanisms that compose and maintain brain health, and understanding how their dysregulation may lead to AD-related pathological changes. AD is a complex neurodegenerative disorder, characterized by the accumulation of amyloid-beta (A*β*) plaques and tau protein tangles, as well as neuronal losses, leading to cognitive decline and memory impairment^2–4^. Recent therapeutic strategies targeting A*β* have been successful in reducing plaque accumulation but are not sufficient to stop the progression of neurodegeneration ^5^. This limitation underscores the need for a more comprehensive strategy that not only understands and targets A*β* and tau pathologies but also other contributing factors known to play a role in the inception and progression of the disease ^6;7^. Besides age, the apolipoprotein E (*APOE*) gene is the most well-established *non-modifiable* risk factor, with the *APOE4* allele conferring a higher risk of both onset and progression of AD^8^. The presence of *APOE4* has been associated with an increased and earlier pathological burden, but the exact mechanisms by which it contributes to disease progression remain an active area of research ^9^. Meta-analyses of epidemiological studies have determined *modifiable* risk factors at the population level (less education, uncorrected hearing loss, hypertension, smoking, obesity, depression, physical inactivity, diabetes, excessive alcohol, traumatic brain injury, air pollution, social isolation, high cholesterol, and uncorrected vision loss) ^10;11^. Livingston and colleagues estimate that up to 45% of dementia cases worldwide could be attributable to exposure to these modifiable factors across the life course ^10;11^. Yet, the heterogeneity observed in brain aging and in the progression toward pathological states between individuals highlights the importance of understanding the interplay between modifiable and non-modifiable risk factors, and the need to develop mechanistic models that can characterize trajectories of brain aging and the emergence of pathological states. This will naturally lead to individualized interventions ^12^ that are our best hope of contributing to a decrease in age-adjusted incidence. Hence, it seems crucial to adopt and develop a larger framework to understand the complexity of AD, including not only amyloid/tau accumulation and neuronal losses, but also neuronal, metabolic, and cerebrovascular dysfunctions, alongside neuroinflammation. Although recent brain health frameworks have emphasized the integration of multi-scale biological processes ^13^, implementing such systems-level approaches experimentally remains challenging due to the large number of variables involved and the ethical and logistical issues related to conducting large-scale trials ^14^. Hence, although these factors have been extensively studied individually, few attempts have been made to integrate them into a holistic framework that can account for their combined effects on brain aging trajectories and biological dysregulation. Mathematical models provide a powerful alternative and systematic framework for solving this issue, by enabling the study of the interplay of multiple biological factors under controlled and reproducible conditions. These models consist of computer programs representing a complex nonlinear system of mathematical equations, derived from prior domain knowledge but validated using simulated and/or real data. In this framework, causal structure is specified a priori within the system of equations, while predictive performance serves as the criterion for validation. Our team has proposed such a mathematical model that integrates interactions across multiple scales, from nanoscale protein dynamics through microscale cellular processes, simulating a 50-years trajectory of brain health ^15^. Importantly, this model was developed as a mechanistic representation of healthy brain aging. The biological processes represented in the model are present throughout normal aging, and AD-related changes may emerge as a consequence of the interactions among these processes when model variables reach pathological levels. At this stage, the model does not explicitly incorporate disease-specific insults or AD risk factors. Thus, its primary purpose is to characterize biological trajectories during healthy aging, while providing a framework for investigating how pathological states may emerge from altered aging processes. Describing the temporal evolution of factors important for brain health throughout aging, the model is composed of 19 different ordinary differential equations (ODE) and 75 parameters. The governing equations are provided in Appendix A, while the full list of optimized parameters and their biological interpretations are presented in Appendix B. Each parameter represents a biologically significant entity or process related to the production, aggregation, and clearance of A*β* and tau protein tangles; neuronal and astrocytic survival and death; activation and polarization of astrocytes, microglia, and macrophages; and the dynamics of inflammatory cytokines such as TNF-*α*, IL-10, TGF-*β* and MCP-1. Many parameters also vary with sex and *APOE* status, allowing the model to test their influence on variability in these aging-related trajectories. Importantly, the model does not assume that all individuals follow the same aging trajectory. Rather, inter-individual heterogeneity can emerge from differences in model parameter values and distributions, such that distinct parameter vectors generate individual-specific trajectories while preserving the same underlying biological mechanisms ^15^. A systematic review of mathematical modeling studies in this field highlighted that verification and validation are often overlooked ^16^. Motivated by this gap, we previously performed single and pairwise parameter perturbations of the parameters of the ^15^ model to study local sensitivities ^17^, primarily investigating how different biological parameters influence neuronal decline and brain aging trajectories. These analyses highlighted the importance of investigating interactions among biological parameters and motivated the need for a more comprehensive multi-parametric sensitivity analysis to better characterize the contribution of biological processes within the model.

In this work, our objective was to further characterize and validate the Chamberland et al.’s mechanistic mathematical model of healthy brain aging. Specifically, we aimed to (i) identify the biological parameters that most strongly influence model outcomes through multi-parametric sensitivity analysis, (ii) calibrate the model using observed biological outcomes from cognitively normal (CN) participants in the Alzheimer’s Disease Neuroimaging Initiative (ADNI), (iii) internally validate the calibrated model in an independent cognitively normal ADNI cohort, and (iv) externally validate its generalizability using an independent healthy population from the UK Biobank cohort. Finally, through variance decomposition analyses in simulated populations, we quantified the contribution of key biological factors, including sex and *APOE* status, to variability in model outcomes. Together, these analyses were expected to provide a comprehensive characterization of the model, assess its ability to reproduce observed biological outcomes and generalize across independent populations, and provide insight into the biological sources of inter-individual variability.

This work provides a biologically interpretable framework for studying the complex interactions underlying healthy brain aging and the potential emergence of AD-related pathological changes. By integrating multiple biological processes within a mechanistic model, this approach may contribute to a better understanding of how individual differences shape brain aging trajectories and provide a foundation for future studies investigating how the incorporation of disease-related risk factors and pathological insults may change these trajectories. In the longer term, such a framework may contribute to the development of more targeted strategies for maintaining brain health and reducing the burden of age-related neurodegenerative disease.

## Methods

### Ethics approval

Our study was conducted with the approval of the Institutional Review Board of Neuroscience and Mental Health of the Centre Intégŕe Universitaire de Santé et Services Sociaux de la Capitale Nationale (CER-CIUSSS-CN NSM #2023–2773) for the project. This research also uses secondary data obtained from the UK Biobank (Application ID #85063) and the Alzheimer’s Disease Neuroimaging Initiative database (Data Use Agreement revised 09/25).

### Data sources

Data used for training and internal validation were obtained from cognitively normal (CN) participants in the Alzheimer’s Disease Neuroimaging Initiative (ADNI) database, a publicly available research resource. The ADNI was launched in 2003 as a public-private partnership led by Principal Investigator Michael W. Weiner, MD. Its primary goal is to test whether serial MRI, PET, other biological markers, and clinical and neuropsychological assessments can measure the progression of mild cognitive impairment (MCI) and early Alzheimer’s disease (AD). Up-to-date information is available at https://adni-info.org.

Furthermore, this study used data from the UK Biobank (UKBB), a large publicly available population-based biomedical resource that contains health, genetic and imaging information from participants across the United Kingdom. Data from cognitively normal (CN) participants in the UK Biobank were used for external validation of the model. Detailed information on participant recruitment, data acquisition, and quality control procedures is available in UK Biobank website.

### Multi parametric sensitivity analysis

To assess the sensitivity of our model to concurrent changes in more than two parameters, we generated a virtual population of Monte Carlo samples, where each sample was represented by one instance of the model with randomly perturbed parameter values. This approach allowed us to capture individual variability, explore how different parameter combinations influenced model outcomes, and identify which factors drove neurodegeneration the most. We assumed for this purpose that every parameter belonged to a normal and independent distribution, regardless of *APOE* or sex status. However, sex and *APOE* status were incorporated using baseline parameters values, so that *APOE+* and *APOE−* individuals, and women and men, started with different baseline values reflecting their biological differences. The analysis was performed separately for each subgroup of sex and *APOE* status (*Women/Men*, *APOE−/APOE*+).

#### Model outcomes

Given that there are 19 model outcomes to test (each ODE results in a distinct model outcome), and that many are not directly measurable in vivo, we focused on a representative subset of outcomes that are sufficient to evaluate model coherence and allow comparison with clinical observations. Model validation generally relies on evaluating key quantities of interest that are biologically relevant and experimentally measurable rather than testing every simulated variable. Therefore, we selected three outcomes that represent the principal biological processes included in the model and relevant to brain aging and Alzheimer’s disease: (a) the density of A*β* plaques, as measured by A*β* positron emission tomography (PET), representing amyloid accumulation; (b) the levels of total tau, as measured in the cerebrospinal fluid, representing tau pathology and neuronal injury; and (c) neuronal density, expressed as estimated neuronal mass (g) using brain volume extracted from anatomical magnetic resonance imaging (MRI), representing neurodegeneration and neuronal loss.

For model calibration and validation, these three outcomes were selected because they are measurable in the ADNI dataset and provide a direct link between model predictions and observed biological changes in Alzheimer’s disease. For each simulation, we therefore recorded the values of A*β*, tau, and N, allowing the correlation between the parameter values and biomarker outcomes to be evaluated.

#### Using truncated normal distributions

When dealing with parameter values, especially in cases where extreme values could lead to unrealistic or undesirable outcomes, using a truncated normal distribution can be beneficial. We used heuristics to ensure that all parameter perturbations remained within biologically plausible ranges and to prevent unrealistic model behavior during simulations. Overall, parameters were allowed up to 50% deviation above or below the baseline parameter value, which was large enough to explore meaningful effects on model outcomes while still maintaining biological realism.

#### Pseudo-algorithm

Our approach can be summarized by the following pseudo-algorithm:

1. Fix the number of Monte Carlo samples (*N*_sample_ = 2000). This sample size was chosen to balance computational cost with statistical reliability: with 2000 Monte Carlo samples, the standard error of correlation estimates is sufficiently small (around 2%), ensuring stable and reliable sensitivity results.
2. Generate a set of perturbed parameters by choosing relative changes for all model parameters from independent Gaussian distributions, with mean 0 and standard deviation 0.1 (corresponding to 10% of the original value). Each relative change (Δ) was truncated to be within the range [-0.5, 0.5] to ensure realistic variations, corresponding to a maximum ±50% change relative to the original parameter. The new parameter values were then calculated as follows:

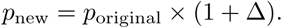 Specifically *N*_sample_ = 2000 random parameter vectors were generated, each consisting of 75 perturbed parameter values.
3. Run the model with these perturbed parameters to obtain output variables. Specifically, focus on *N*_sample_ values of *Aβ*, tau, and *N* at 80 years of age to evaluate the long-term behavior of the model and quantify the influence of parameter perturbations on the model outputs over the adult life span.
4. Calculate the correlation between parameter perturbations and model outcomes (A*β*, tau and neuronal density).

This Monte Carlo simulation approach, which samples relative parameter perturbations from a normal distribution to determine their influence on biomarker levels, allowed us to quantify the strength and direction of the relationship between parameters and outcomes of interest. Prior to correlation analysis, the normality of the simulated biomarker outputs was assessed using the Shapiro–Wilk test. Because the outputs did not satisfy the assumption of normality, Spearman rank correlation was used to evaluate monotonic relationships between parameter values and biomarker outcomes. Differences in Spearman correlation coefficients across sex and *APOE* subgroups were assessed using permutation tests with 10,000 random permutations. The number of permutations was chosen to provide stable p-value estimates while maintaining computational efficiency.

### Quantitative validation of outcomes against cohorts data

#### Dataset partition

For calibration purposes, we selected 765 participants from ADNI, including individuals with cognitively normal (CN), mild cognitive impairment (MCI), and Alzheimer’s disease (AD). Although more participants were available, we included only those with complete follow-up information, ensuring that all outcome measures were fully observed. CN participants were randomly split into a training set (60%), a validation set (20%) and an independent test set (20%) for exploratory evaluation. All MCI and AD participants were assigned to the independent test set. For quantitative validation purposes, we selected 35,899 CN participants from the UKBB with available brain segmentation volume and complete covariate information. These participants served as an independent cohort to assess the generalizability of neuronal density (N) predictions obtained from parameters optimized using ADNI. Only CN participants were included from UKBB because the objective of this external validation was to evaluate neuronal density predictions in a large healthy aging population. The demographic characteristics, including sex distribution across ADNI diagnostic groups and the UKBB cohort, are summarized in Table 1

**Table 1:** Demographic and clinical characteristics of study participants from ADNI and UKBB group.

| Characteristic | CN (n=263) | MCI (n=376) | AD (n=126) | CN (n=35899) |
| --- | --- | --- | --- | --- |
| **Dataset** | ADNI | ADNI | ADNI | UKBB |
| Male ($n$) | 121 | 209 | 76 | 16970 |
| Female ($n$) | 142 | 167 | 50 | 18929 |
| Sex (% Male) | 46.0% | 55.6% | 60.3% | 47.3% |
| Age (years) | $74.5 \pm 6.9$ | $72.3 \pm 7.7$ | $74.7 \pm 8.2$ | $55.0 \pm 7.5$ |
| BrainSegVol(mm<sup>3</sup>) | $1,038,637 \pm 104,995$ | $1,054,501 \pm 104,759$ | $1,001,577 \pm 118,108$ | — |
| Centiloid | $19.2 \pm 36.7$ | $39.1 \pm 48.0$ | $78.5 \pm 47.9$ | — |
| p<sub>tau</sub>CSF (pg/mL) | $22.3 \pm 9.4$ | $25.3 \pm 11.9$ | $34.0 \pm 12.6$ | — |

\*Values are expressed as mean ± standard deviation. CN: cognitively normal; MCI: mild cognitive impairment; AD: Alzheimer’s disease; ADNI: Alzheimer’s Disease Neuroimaging Initiative; UKBB: UK Biobank. Note: “—” indicates data not applicable.

#### Model calibration strategy

The calibration procedure was carried out exclusively on the ADNI training set until convergence of the loss function was achieved. The ADNI validation set was then used to evaluate the performance of the calibrated parameters, without contributing to the parameter calibration or affecting the convergence in any way. The ADNI test set was then used for internal validation (i.e. within same study), and the UKBB test set for external validation (i.e. generalization to another study).

To calibrate the model parameters, we used the model to generate predictions for each ADNI CN training set participant by initializing the model at 30 years of age and running until the age at which the participants received their first MRI. The comparison of outcomes was made on that first scan. The model was instantiated using demographic (sex) and genetic (*APOE4* allele) status. Furthermore, we included baseline neuronal volume (*N*_0_) as an initial condition, derived from brainvol_30_ (30 years), which was calculated from the total intracranial volume (eTIV) and the sex of the participants using a regression equation adapted from ^18^

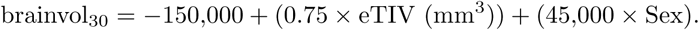

The assumption being, of course, that while brain volume varies, intracranial volume does not change with time. The baseline neuronal volume *N*_0_ was individualized for each subject to ensure physiologically realistic initial conditions.

The ODEs were solved using Python’s integrate.solve_ivp function of SciPy package ^19^ with the BDF (backward differentiation formula) method, which is suitable for stiff systems. The system is considered stiff because it contains processes that evolve on very different timescales, for example, hours for molecular events, days to months for cellular processes, and decades for system-level changes such as brain atrophy. In all simulations, the numerical solver was run with an absolute tolerance of atol = 1 *×* 10^−10^ and a relative tolerance of rtol = 1 *×* 10^−8^ to ensure numerical integration of the ODE ^20^.

The model parameters were calibrated by minimizing the weighted loss function across our main outcomes of interest, namely A*β*, tau and neuronal density. The loss function consisted of the mean squared error across subjects using the logarithmic error for A*β* and tau and relative errors for neuronal density. Parameter calibration was performed using the L-BFGS-B (Limited-memory Broyden–Fletcher–Goldfarb–Shanno with Bound constraints) algorithm implemented in SciPy, by varying the 30 most influential parameters identified through local sensitivity analysis in our previous work ^17^ within physiologically plausible ranges.

#### Internal validation

Once calibrated, the model was used to generate predictions that were used for internal validation, i.e., within the same source of data (ADNI). To this end, we focus on the same three outcomes of interest: N, A*β*, and tau. The prediction accuracy for each outcome was quantified using the mean absolute relative error (MARE), visualized using box plots, and assessed by comparing predicted and observed values using scatter plots. All analyses were performed separately for women and men. Statistical differences in prediction errors before and after calibration were assessed using the Wilcoxon signed-rank test, as the paired error distributions did not satisfy normality assumptions. To assess the consistency of model performance between the training and validation cohorts, the distributions of prediction errors after optimization were compared using the two-sample Kolmogorov–Smirnov test. Correlations between predicted and observed outcomes were assessed using Spearman rank correlation. Normality was evaluated using the Shapiro–Wilk test, and Spearman correlation was selected because several variables did not satisfy the normality assumption required for Pearson correlation.

For comparison with MRI-derived measurements, the predicted neuronal density *N* at the time of MRI was multiplied by the individual’s baseline brain volume at 30 years of age (brainvol30) to obtain the predicted neuronal mass in grams, which was compared to the actual observed neuronal mass (*N*_real_). The prediction accuracy was visualized using box plots showing the mean absolute relative error MARE (Mean Absolute Relative Error) before and after parameter calibration and scatter plots illustrating the correlation between predicted and observed neuronal mass.

For the validation of the dynamics of A*β*, we used ADNI’s standardized centiloid values as a proxy for the accumulation of brain A*β* to evaluate the accuracy of our model predictions. The predicted amyloid beta (A*β*_predicted_) at the time of (Positron Emission Tomography) PET scan was compared to the centiloid of ADNI data, which serve as standardized measurements of amyloid-beta (A*β*) deposition in the brain. The real amyloid-beta concentration was calculated as

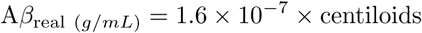

where the conversion factor 1.6 *×* 10^−7^ was derived from ^21^. This evaluation allowed us to assess how closely the model predictions matched the real-world A*β* measurements, providing insight into the model’s performance on external real-world data. The predictive accuracy of A*β* was quantified using MARE, visualized using box plots, and further examined by comparing predicted and observed values.

To validate tau dynamics, cerebrospinal fluid phosphorylated tau measurements (p-tau_CSF_) from the ADNI data set were used as proxies for the real tau concentration (*τ*_real_). The model predictions were directly compared with these measurements to evaluate performance. The predictive performance for tau was measured using the mean absolute relative error (MARE), with box plots showing error distributions and scatter graphs depicting correlations between predicted and observed values.

#### External validation

Once internally validated, the calibrated model was used as-is (with no refitting) to generate predictions for external validation in an independent healthy aging population, using the UKBB cohort. Although the UKBB includes participants with a range of health conditions and clinical characteristics, the present analysis was restricted to cognitively normal (CN) participants to evaluate the generalizability of brain health predictions in an independent healthy aging population. The UKBB cohort includes a large number of cognitively normal participants and represents a broad adult age range, including relatively younger individuals compared with ADNI. The availability of MRI-based brain measurements and demographic information in UKBB makes it suitable for assessing the generalizability of neuronal density (N) predictions. Since UKBB does not contain comparable A*β* and tau measurements as ADNI, these outcomes could not be externally validated in this dataset. Other datasets are being explored for this purpose (e.g., OASIS). A piecemeal strategy for validation will be required for multiple outcomes as not all databases have such populations and measurements available at scale. For each of the 35,899 CN participants, consisting of 18, 929 women (52.7%) and 16,970 men (47.3%) the calibrated model was used to simulate outcomes, running from age 30 to the age of each participant at the scan, using similar individual characteristics (sex, *APOE4* status, brain volume at 30 y.o. from eTIV) as initial conditions. This generated the predicted neuronal density for each participant, which was then compared to the real neuronal density obtained from MRI separately for women and men. In addition, the distribution of the real neuronal density was visualized using histograms and kernel density estimates (KDEs) stratified by sex and *APOE* status, allowing for the assessment of how neuronal density varied with genetic risk. Differences in the distributions of observed neuronal density between *APOE4−* and *APOE4+* groups were statistically assessed separately for women and men using the two-sample Kolmogorov–Smirnov test. The correlation between predicted and MRI-derived neuronal density was assessed using Spearman rank correlation. The normality assumption was evaluated using the Shapiro–Wilk test, and Spearman correlation was used because the variables did not satisfy Pearson correlation assumptions. This approach provided both a quantitative measure of the model accuracy (by correlation) and a qualitative assessment of the distributional patterns of neuronal density in the cohort.

### Proportion of explained variance

Finally, we wanted to qualitatively evaluate the proportion of variance that could be attributed to genetic parameters (e.g. *APOE4* status) when compared to calibrated biophysical parameters. To this end, we developed the following methodology and applied it to the important case of the interaction between *APOE* status and A*β*, *N*, and tau levels. We adopted a similar Monte Carlo approach and generated 2,000 simulated samples, stratified according to *APOE* status: carriers (one or two e4 alleles: e4/e4, e3/e4, e4/e3, e2/e4 or e4/e2) and non-carriers (no e4 allele: e3/e3, e2/e3, e3/e2, or e2/e2). In our simulations, 40% of the individuals were assigned as *APOE4* carriers and 60% as non-carriers ^22;23^. By incorporating this stratification into Monte Carlo population-based simulations, we aimed to better capture the heterogeneity in disease progression observed in real world populations. From these simulations, we computed the proportion of variance explained by both *APOE* status and the optimized model parameters, enabling us to quantify their respective contributions to variability in disease trajectories within the simulated cohort.

## Results

### Multi-parametric sensitivity analysis

The correlation graphs in (Figure 1) illustrate the relationships between various parameters and our three main outcomes (*Aβ*, *N* and tau) in our multi-parametric sensitivity analysis. Spearman rank correlation coefficients and corresponding p-values were calculated to quantify the associations between model parameters and biomarker outputs. The corresponding p-values for all correlation analyses were below 0.001. To identify the most influential parameters, parameters in each plot are ranked in descending order based on the average of their absolute correlation coefficients across all four cases (Sex (Men, Women) and *APOE4* status (present, not present)).

**Figure 1:**
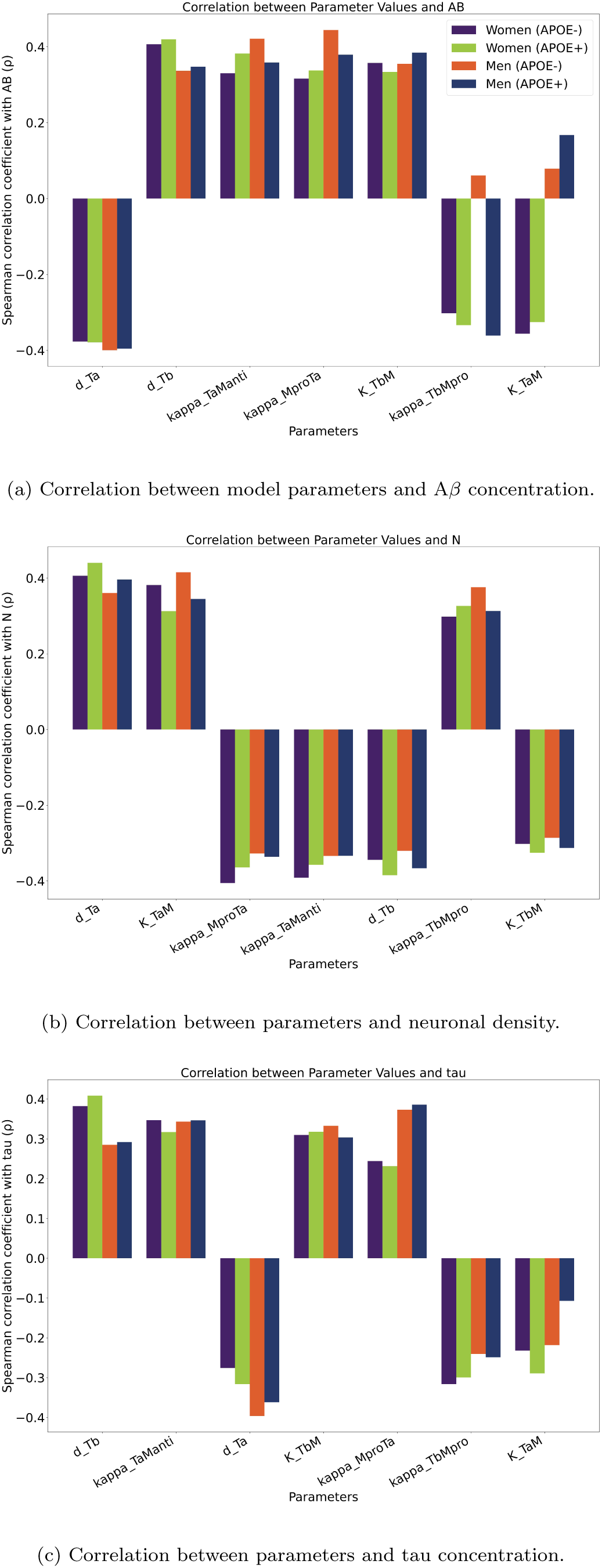
Spearman correlation coefficients (*ρ*) between model parameters and biomarkers at age 80 years across different demographic groups (men and women, *APOE+* and *APOE-*). The outcomes considered are **(a)** A*β*, **(b)** *N* and **(c)** tau. The virtual population was generated with *σ* = 0.10 and *N*_sample_ = 2000. All reported correlations were statistically significant (*p <* 0.001).

Figure 1 reveals that many of the parameters highly correlated with outcome values were also identified in our single-and pair-parameter sensitivity analysis ^17^.

Sorting parameters by average absolute correlation identified *d*_Ta_ (TNF-*α* decay) and *d_T b_* (TGF-*β* decay) among the parameters showing the strongest associations across the three outcomes. For example, the parameter *d*_Tb_ (decay rate of *TGF* -*β*) shows a consistent positive correlation with *Aβ* and tau, whereas *d*_Ta_ showed a negative correlation with *Aβ* and *tau*. Notably, *d*_Ta_ showed the strongest positive correlate of neuronal density (N).

We further observed that the parameter *K_T bM_* is positively correlated with A*β* and tau concentrations, while it is negatively correlated with N. This parameter represents the sensitivity of macroglia activation to the concentration of TGF-*β*.

Although the magnitude of correlations was generally similar across groups, some parameter biomarker associations differed significantly between specific sex/*APOE* subgroups. In the *Aβ* plot, the parameters *K_T aM_* and kappa*_TbMpro_* showed distinct difference between women *APOE-*and men *APOE-*. *K_T aM_*displayed a negative correlation in women *APOE-*(*ρ* = *−*0.356) compared with a weak positive correlation in men *APOE-*(*ρ* = 0.079). Similarly, kappa*_TbMpro_* showed a negative correlation in women *APOE-*(*ρ* = *−*0.302) compared with a weak positive correlation in men *APOE-*(*ρ* = 0.061). These difference were statistically significant based on permutation testing (*p <* 0.001). These differences indicate that the associations between specific model parameters and A*β* differed between women *APOE−* and men *APOE−*. Overall, the correlation analysis identified the parameters most strongly associated with model outputs and characterized their relationships with A*β*, tau, and neuronal density. The abbreviations of the model parameters presented in this analysis are summarized in Table 2.

**Table 2:** Description of model parameter abbreviations used in the figures.

| Parameter | Description |
| --- | --- |
| $d_{Ta}$ | Degradation rate of Tumor Necrosis factor alpha (TNF-α). |
| $\text{kappa}_{TaManti}$ | Maximal conversion rate of anti-inflammatory microglia to proinflammatory under TNF-alpha signaling. |
| $d_{Tb}$ | Degradation rate of Transforming Growth Factor beta (TGF-β). |
| $K_{TbM}$ | Concentration of TGF-β at which the conversion of pro-inflammatory microglia $M_{pro}$ to anti-inflammatory microglia $M_{anti}$ is half-maximal. |
| $\text{kappa}_{MproTa}$ | Production rate of TNF-α by pro-inflammatory microglia ($M_{pro}$). |
| $\text{kappa}_{TbMpro}$ | Maximal conversion rate of pro-inflammatory microglia ($M_{pro}$) to anti-inflammatory microglia ($M_{anti}$) under TGF-β signaling. |
| $K_{TaM}$ | Concentration of Tumor Necrosis factor alpha (TNF-α) at which the conversion of anti-inflammatory microglia $M_{anti}$ to pro-inflammatory microglia $M_{pro}$ is half-maximal. |
| $d_{MantiABpo}$ | Degradation rate of extracellular amyloid-beta42 plaque by anti-inflammatory microglia ($M_{pro}$). |
| $\text{lambda}_{ABmo}$ | Creation rate of amyloid-beta monomer outside (without APOE4 allele). |
| $\text{Ins}_0$ | Normal concentration of insulin, sex dependent. Correspond to the brain concentration at 30 years old. |
| $\text{kappa}_{ABmoABoo}$ | Conversion rate of extracellular amyloid-beta monomer to extracellular amyloid-beta oligomer. |
| $d_{ABoo}$ | Degradation rate of extracellular amyloid-beta42 oligomer. |
| $d_{Fi}$ | Degradation rate of intracellular NFT. |
| $\text{kappa}_{MhatproTa}$ | Production rate of TNF-alpha by proinflammatory macrophages ($hat{M}_{pro}$) |
| $\text{lambda}_{Gtau}$ | production rate of soluble tau (Gtau). |
| $\text{kappa}_{tauFi}$ | Conversion rate of tau in NFT. |
| $n$ | Sigmoid function coefficient. |
| $\text{kappa}_{TaMhatanti}$ | Maximal conversion rate of anti-inflammatory macrophage to proinflammatory under $TNF - \alpha$ signaling. |
| $d_{tau}$ | Degradation and un-hyperphosphorylation rate of intracellular tau proteins. |
| $\text{delta}_{Apmo}$ | This constant quantifies the impact of the APOE4 gene on the conversion rate of extracellular amyloid-beta monomer to extracellular amyloid-beta oligomers. |
| $\text{delta}_{Apdp}$ | This constant quantifies the impact of the APOE4 gene on the degradation rate of amyloid-beta42 plaque outside by anti-inflammatory macrophages and microglia. |
| $d_{TaN}$ | Maximal death rate of neurons induced by $T_\alpha$ (TNF-alpha). |

### Quantitative validation of predicted outcomes against cohorts data

#### Demographic and clinical characteristics of study cohorts

The demographic and clinical composition of the study participants, which comprises both the ADNI (*N* = 765) and UKBB (*n* = 35, 899) cohorts, is detailed in Table 1. Participants are categorized by diagnostic group (CN, MCI, and AD). The ADNI cohort was composed of 263 CN, 376 MCI and 126 AD participants. Sex distribution vaied across these groups; while the CN group included 142 women (54.0%) and 121 men (46.0%), the MCI group included 167 women (44.4%) and 209 men (55.6%), and the AD group included 50 women (39.7%) and 76 men (60.3%). The mean age remained consistent across all ADNI groups, falling within the 74–75 year range. This overlapping age distribution supports demographic comparability and minimizes age as a confounding factor for model validation. The age at first MRI ranged from 59 to 93 years, with an average simulation duration of 44 years (from start point at 30 y.o.).

The key biomarkers in the ADNI cohort reflected expected clinical patterns: BrainSegVol was significantly lower in the AD group than in both CN and MCI, consistent with brain atrophy; Centiloid values increase progressively from CN to MCI to AD, consistent with rising amyloid burden; and p-tau*_CSF_* concentrations similarly increased with disease severity. To statistically assess these differences, normality was first evaluated using the Shapiro–Wilk test, which indicated non-normal distributions for these biomarkers. Therefore, the nonparametric Kruskal–Wallis test was used to compare biomarker distributions across CN, MCI, and AD groups. Significant group differences were observed for BrainSegVol (H = 20.329, *p <* 0.001), Centiloid values (H = 108.723, *p <* 0.001), and p-tauCSF concentrations (H = 81.575, *p <* 0.001). Dunn’s post-hoc tests with Bonferroni correction showed that BrainSegVol was significantly lower in the AD group compared with both CN (p = 0.0096) and MCI (*p <* 0.001) whereas the difference between CN and MCI was not significant (*p* = 0.224). For Centiloid values, all pairwise comparisons (CN vs. MCI, CN vs. AD, and MCI vs. AD) were significant (all *p <* 0.001), confirming the progressive increase across disease stages. Similarly, p-tauCSF concentrations differed significantly across all pairwise comparisons (CN vs. MCI, *p* = 0.0084; CN vs. AD and MCI vs. AD, both *p <* 0.001). In contrast, the UKBB cohort provides a massive reference population of 35,899 CN individuals. This cohort is characterized by a slightly higher proportion of females (52.7%, *n* = 18, 929) compared to males (47.3%, *n* = 16, 970). The mean age of the UKBB cohort is 55.0 *±* 7.5 years, representing a middle-aged population, with an average model run time 34 years from start age (30 years) to age at scan.

The UKBB cohort was used exclusively for external validation of neuronal density predictions, allowing assessment of the generalizability of the calibrated model in a large, independent cognitively normal population.

#### Effect of optimization tolerance on model performance

We tested different ftol optimization tolerance levels (function tolerance) to assess their impact on computational time and prediction accuracy. Across all values tested, the optimizer converged to essentially identical prediction errors for A*β*, tau, and neuronal density, while tighter tolerances substantially increased computational time. This indicates that moderate tolerance values (e.g., ftol = 10^−10^ or 10^−12^) were sufficient for efficient parameter calibration without loss of accuracy.

#### Internal validation of neuronal mass

In (Figure 2a) the sex-specific Spearman correlation analysis reveals strong positive relationships between the real and predicted values for neuron mass (*N*) in the Cognitively Normal (CN) cohort. Specifically, the model yielded a correlation coefficient of *ρ* = 0.69, (*p <* 0.001, *n* = 112) for women and *ρ* = 0.72, (*p <* 0.001, *n* = 99) for men, indicating that the model demonstrated strong correlations between predicted and observed neuronal mass in both sexes.

**Figure 2:**
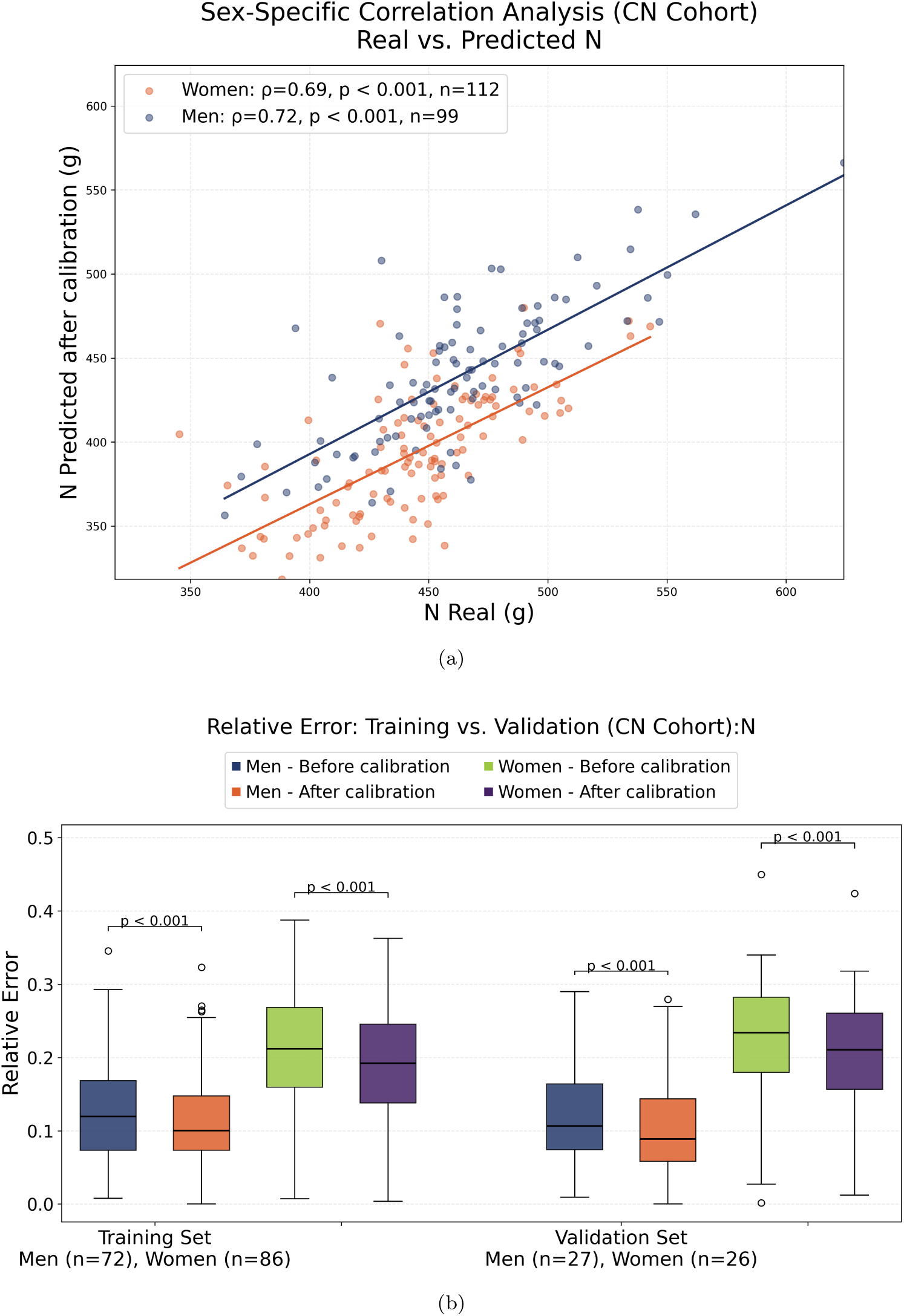
**(a)** Correlation between real and predicted neuron mass after calibration in the CN cohort, stratified by sex (women: *ρ* = 0.69; men: *ρ* = 0.72; both *p <* 0.001). **(b)** Relative error distributions before and after calibration for the training (*n* = 158) and validation (*n* = 53) sets. Calibration significantly reduced relative error across all sub-groups (*p <* 0.001, Wilcoxon signed-rank test).

(Figure 2b) shows the distribution of relative errors before and after calibration across both the training set (Men: *n* = 72, Women: *n* = 86) and the validation set (Men: *n* = 27, Women: *n* = 26). In the training set, calibration reduced the mean relative error from approximately 0.12 to 0.10 for men and from 0.21 to 0.19 for women. In the validation set, the mean relative error dropped from 0.11 to 0.09 for men and from 0.23 to 0.21 for women. Wilcoxon signed-rank tests showed statistically significant differences between pre-and post-calibration relative errors across all subgroups (*p <* 0.001 for each comparison).

#### Internal validation of predicted Aβ pathology

To evaluate the predictive performance for A*β* concentration across sex groups within the cognitively normal (CN) cohort, a correlation analysis was conducted between real and predicted A*β* levels (Figure 3a). A statistically significant, weak positive correlation was observed in men (*ρ* = 0.29*, p* = 0.004*, n* = 99), whereas the correlation in women was weaker and not statistically significant (*ρ* = 0.14*, p* = 0.137*, n* = 112). Model predictions exhibited a clustered distribution across both groups, suggesting limited sensitivity to extreme individual variability in A*β* levels.

**Figure 3:**
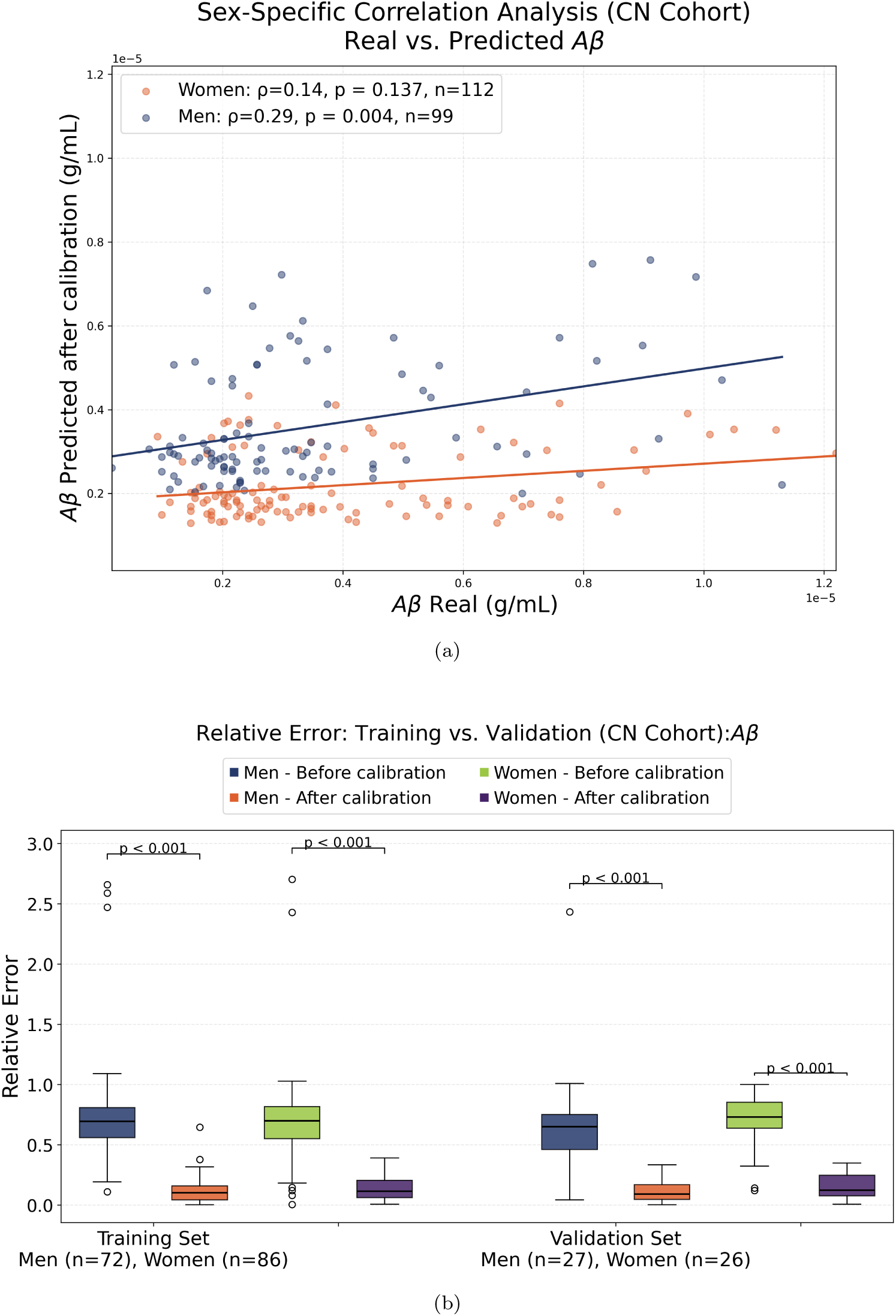
**(a)** The scatter plot illustrates the relationship between real A*β* deposition, measured in centiloids, and the predicted A*β* values generated by the model, for 211 cognitively normal (CN) individuals aged 59–93. **(b)** Relative Error for training (*n* = 158) and validation (*n* = 53) sets. Calibration reduced the magnitude and variability of prediction errors, improving model performance. All participants were retained in the analysis; apparent extreme values represent larger prediction errors rather than excluded observations

In (Figure 3b), we looked at the relative prediction errors before and after calibration across our training (men: n = 72, women: n = 86) and validation (men: n = 27, women: n = 26) sets. Prior to calibration, relative errors were substantially elevated and displayed high variability. Following calibration, relative errors decreased significantly across all subgroups (*p <* 0.001 for all comparisons, Wilcoxon signed-rank test). Specifically, in the training set, mean relative errors dropped from approximately 0.70 to 0.10 in men and from 0.70 to 0.12 in women. A consistent reduction was observed in the validation set, where relative errors decreased from 0.65 to 0.10 in men and from 0.75 to 0.15 in women.

#### Internal validation of tau concentrations

In (Figure 4a), sex-specific Spearman correlation analysis showed a moderate positive correlation between observed and predicted tau concentrations in men (*ρ* = 0.39, *p <* 0.001, *n* = 99). In women, a weak positive correlation was observed that did not reach statistical significance (*ρ* = 0.06, *p* = 0.556, *n* = 112).

**Figure 4:**
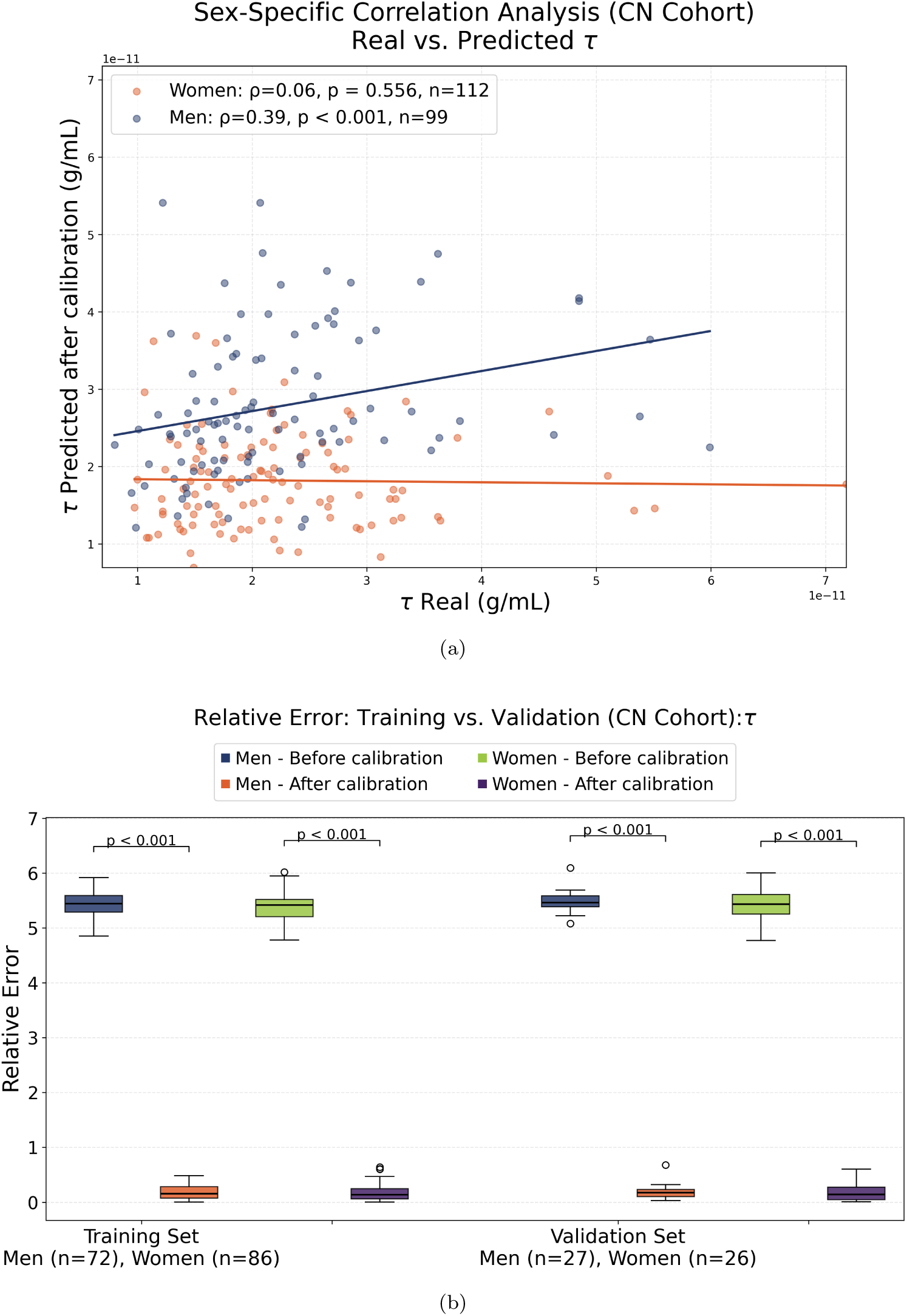
**(a)** The scatter plot illustrates the relationship between real tau, measured in ptau*_CSF_*, and the predicted tau concentrations generated by the model, for 211 cognitively normal (CN) individuals aged 59–93. **(b)** Boxplot of mean relative error for training (*n* = 158) and validation (*n* = 53) sets. Calibration reduced the magnitude and variability of prediction errors in both sexes.

(Figure 4b), illustrates the relative prediction errors before and after calibration, evaluated separately for men and women across the training set (Men: *n* = 72, Women: *n* = 86) and the validation set (Men: *n* = 27, Women: *n* = 26). Prior to calibration, relative errors were exceptionally high and variable across both sexes. Following calibration, mean relative errors decreased significantly in all subgroups (*p <* 0.001 for all comparisons, Wilcoxon signed-rank test). In the training set, the mean relative errors dropped from approximately 5.4 to 0.20 in men and from 5.4 to 0.20 in women. Similarly, in the validation set, mean relative errors decreased from 5.5 to 0.20 for men and from 5.4 to 0.20 for women.

#### External validation of neuronal density

(Figure 5a) shows the Spearman correlation coefficient between the real and predicted neuronal density in the UKBB cohort. In men, the correlation was *ρ* = 0.86 (*p <* 0.001), while in women it was slightly higher at *ρ* = 0.87 (*p <* 0.001).

**Figure 5:**
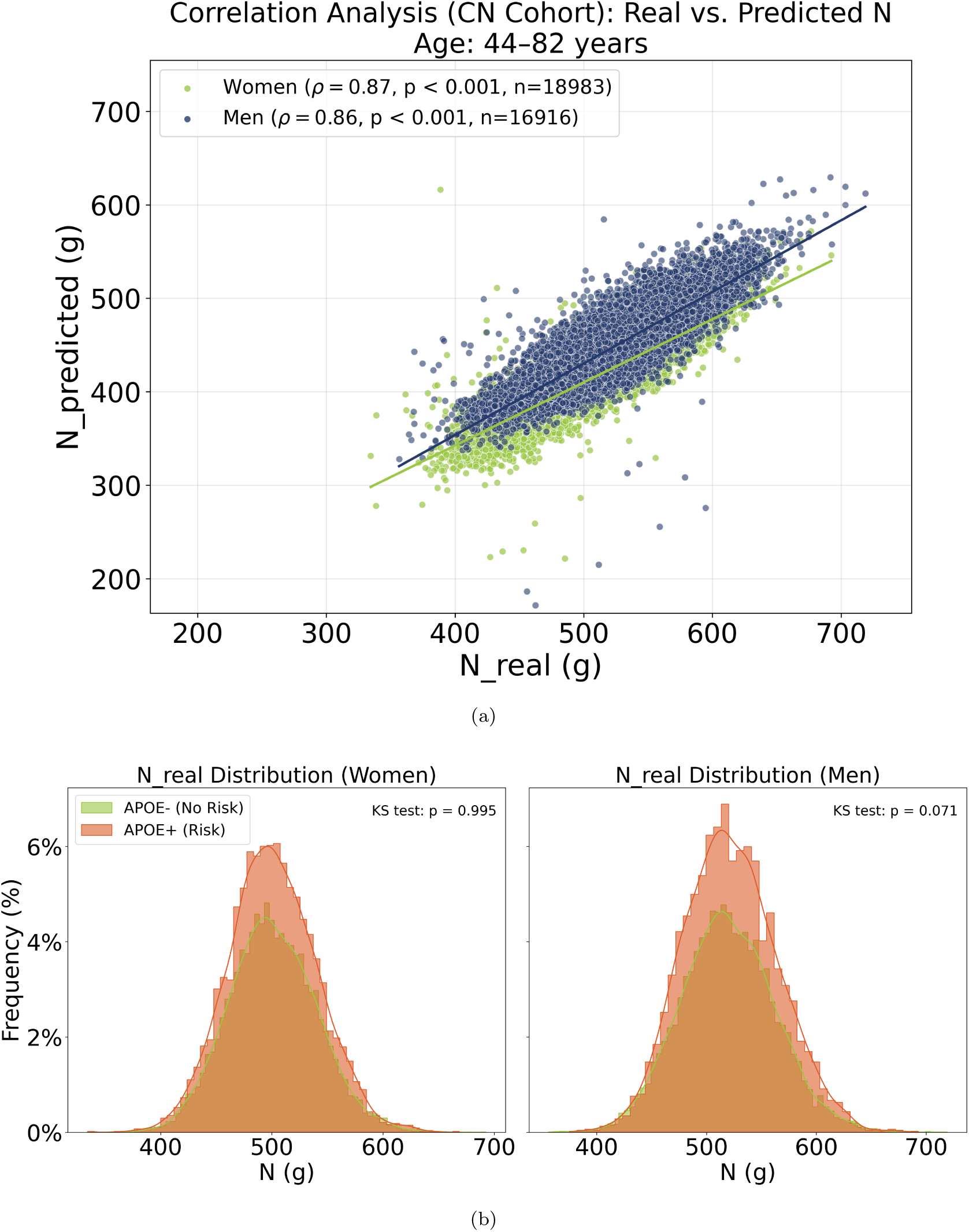
**(a)** Scatter plot showing the relationship between observed neuronal mass (*N*_real_), derived from brain segmentation volume, and the predicted neuronal mass values generated by the model for 35,899 individuals aged 44–82 years. **(b)** Distribution of observed neuronal density in the UKBB cohort according to *APOE4* status. Histograms and kernel density estimates (KDE) illustrate the distributions for *APOE4−* and *APOE4+* groups separately in women and men. Two-sample Kolmogorov–Smirnov tests showed no statistically significant differences between *APOE4−* and *APOE4+* distributions in women and men.

In (Figure 5b), histograms and kernel density estimates (KDE) illustrate the frequency distribution of the values observed *N* (*g*) in all subgroups. Two-sample Kolmogorov–Smirnov tests showed no statistically significant differences in the distributions of observed neuronal density between the “No Risk” (*APOE4-*) and “Risk” (*APOE4+*) groups in either women (*D* = 0.0067, *p* = 0.995) or men (*D* = 0.0223, *p* = 0.071).

### Proportion of explained variance

The proportion of variance explained by *APOE4* in amyloid-beta (A*β*), neuronal density (N), and tau was analyzed separately for men and women using the optimized model parameters (Figure 6).

**Figure 6:**
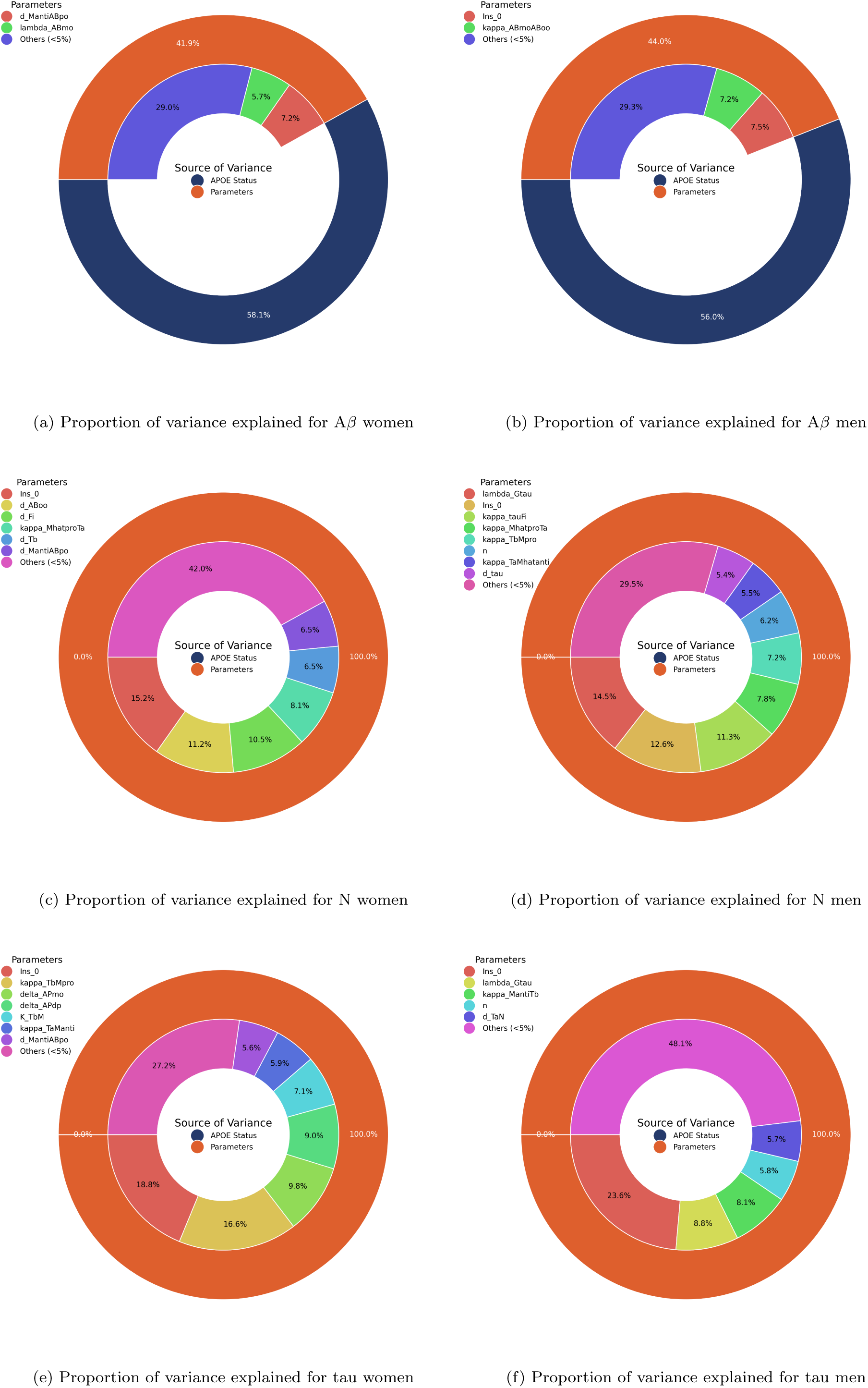
Variance explained for A*β*, neuronal densit_2_y_2_ (N), and tau by *APOE* status and optimized model parameters across sex groups. Only parameters explaining *>* 5% of variance are shown.

For A*β* (Figures 6a and 6b), *APOE* status explains a substantial proportion of the variance in women 58.1%, while the remaining 41.9% is attributed to other model parameters. In contrast, in men, *APOE* explains a proportion of the variance of 56.0%, compared to 44.0% explained by the other parameters.

For neuronal density (Figures 6c and 6d), *APOE* contributes negligibly to variance. In men and women, none of the variance is explained by *APOE* 0%, while the remaining 100% is explained by other parameters.

A similar pattern is observed for tau (Figures 6e and 6f). In women, *APOE* accounts for 0.1% of the variance, with 99.9% explained by other parameters. In men, 100% of the variance is explained by the other parameters, indicating that *APOE* has a negligible effect on tau variability. Overall, these results indicate that, after calibration, *APOE* strongly influences A*β* variability but has minimal impact on neuronal density and tau variability. The abbreviations of the model parameters presented in this analysis are summarized in Table 2.

## Discussion

This study explores the validity of our model of brain health dynamics, as well as the significant influence of key model parameters on processes associated with neurodegenerative processes.

### Multi-parametric analyses

Multi-parametric sensitivity analysis revealed that model dynamics is influenced by multiple interacting parameters rather than a single dominant factor. his supports the multifactorial nature of brain aging, in which several biological processes may collectively shape aging trajectories across sex and *APOE* subgroups. This is consistent with Alzheimer’s disease being a complex condition, where focusing on a single factor such as amyloid-beta has shown limited success ^24;25^. Computational models provide an ideal integrative framework for testing multi-factorial hypotheses in silico ^26–29^. Inflammation-related parameters, particularly those governing TNF-*α* and TGF-*β* dynamics, highlight the potential importance of immune regulation in brain aging. This interpretation is consistent with previous studies linking neuroinflammatory changes with aging and Alzheimer’s disease-related changes.

The influence of glial-related parameters, including *K_T bM_* and kappa*_TbMpro_*, further emphasizes the potential importance of glial regulation in shaping brain-aging trajectories. Their associations with multiple model outcomes suggest that changes in glial regulation may contribute to the emergence of neurodegenerative-related features within the model as an emergent property of the system.

Furthermore, differences in parameter-biomarker correlation across sex and *APOE* groups highlight the importance of considering population variability in brain aging models. The observed sex-dependent relationships between specific model parameters and A*β* suggest that biological sex and genetic risk may influence how underlying model processes relate to amyloid accumulation. Incorporating such subgroup variability may therefore be important for capturing individual differences in biomarker trajectories and advancing personalized models of brain aging.

### Pathway of action and parameter influence

Each of these key parameters influences different biological pathways relevant for age-related biomarker dynamics and neuroinflammatory regulation: d*_T b_* represents the degradation rate of TGF-*β* (transforming growth factor beta), which is an important anti-inflammatory cytokine ^30^. Increasing *d_T b_* leads to faster breakdown of TGF-*β*, which can reduce its protective effects. In contrast, decreasing d*_T b_* allows TGF-*β* to remain active longer, supporting anti-inflammatory signaling and promoting neural protection.

kappa*_TbMpro_* denotes the maximal conversion rate of pro-inflammatory microglia (*M_pro_*) to anti-inflammatory microglia (*M_anti_*) under TGF-*β* (transforming growth factor beta) signaling. Increasing this conversion rate can enhance anti-inflammatory responses, reduce tau hyperphosphorylation, and mitigate neuronal damage. In parallel, *K_T aM_* represents the concentration of TNF-*α* for which the conversion of anti-inflammatory microglia (*M*_anti_) back into pro-inflammatory microglia (*M*_pro_) is half-maximal, serving as a critical sensitivity threshold for inflammatory phenotype switching. *K_T bM_* represents the concentration of TGF-*β* for which the conversion of pro-inflammatory microglia (*M_pro_*) into anti-inflammatory microglia (*M_anti_*) is half-maximal, defining the regulatory threshold for inflammation resolution and microglial phenotype transition.

kappa*_MproTa_* represents the rate of TNF-*α* production by pro-inflammatory microglia (*M_pro_*). An increase in kappa*_MproTa_* indicates that an increase in TNF-*α* may promote A*β* accumulation, possibly through enhanced microglial activation or impaired clearance mechanisms. However, it contributes to a decrease in neuron density due to inflammation-induced neurodegeneration. kappa*_TaManti_* governs the anti-inflammatory microglial response (*M_anti_*), promotes tissue repair, reduces tau hyperphosphorylation, and mitigates neuroinflammation.

### Parameters calibration and error reduction

Parameter calibration substantially reduced prediction errors across neuronal density, amyloid beta and tau demonstrating the effectiveness of the calibration procedure in improving agreement between model predictions and the observed data. The reduction in relative errors after calibration across outcomes and sex groups indicates improved prediction stability. Importantly, the calibrated parameters remained biologically plausible, suggesting that calibration refined the model without producing unrealistic results. Although calibration reduced prediction errors across all outcomes, the degree of predictive agreement differed between model outputs, highlighting the varying complexity of the biological processes represented by neuronal density, A*β* and tau.

### Internal and external validation

Consistent performance during internal validation using ADNI (CN) data and external validation using the UK Biobank cognitively normal cohort supports the robustness and generalizability of the model across independent cohorts. It is important to recall that the model is one of *normal* brain health. Contrary to other models ^31;32^, there is no injection of a pathological disruptor (e.g. reactive oxygen species) to “generate” a disease state. Rather, initial parameters were selected based on the literature relating to cognitively healthy trajectories; and herein, optimized using CN individuals. The model therefore is one of the normal brain aging dynamics, where neurodegenerative-related features may emerge as consequences of aging-associated biological processes. Internal validation in the ADNI (CN) cohort showed that neuronal mass had the strongest agreement between predicted and observed values across sex groups. The emergence of A*β* and tau is viewed as a consequence of biological processes associated with aging. So, it is not surprising that the model performs best on neuronal mass; and most poorly on A*β* and tau. This distinction was particularly evident in the sex-specific analyses. Neuronal density showed strong and statistically significant correlations between predicted and observed values in both men and women, whereas A*β* and tau showed weaker correlations, with statistically significant correlations observed in men but not in women. These findings suggest that the current model captures neuronal changes associated with normal aging more effectively than individual variability in A*β* and tau concentrations. Future iterations with pathological disruptors (e.g., cardio-metabolic syndrome) may completely change this picture.

This was further supported by the strong performance in predicting neuronal mass in the UKBB, a cognitively healthy cohort, indicating a strong monotonic relationship between predicted and observed values. The stronger association observed in UKBB compared with the ADNI cohort suggests that parameters optimized in a relatively small clinical dataset can generalize effectively to a much larger population-based cohort. The scatter plot in Figure 5a illustrates that the model captures the overall variation in neuronal density extremely well for CN individuals across decades, maintaining high predictive accuracy between diverse genetic risk profiles. The fact that the correlation was higher than in the ADNI (CN) validation cohort also underlined the effect of age. Essentially, the difference could be related to the approximately 10-years difference in MRI scan years, the UKBB simulations not having this extra time to “diverge” from reality.

### Proportion of explained variance for sex/APOE

We evaluated the proportion of explained variance to quantify the relative contribution of model parameters and *APOE* status to the variability of the outcomes. The results showed that *APOE* status explains more than 50% of the variability in A*β* levels, highlighting its dominant influence on amyloid dynamics in the model. In contrast, the contribution of *APOE* to the variability of neuronal density (N) and tau levels was almost negligible. This suggests that while *APOE* plays a major role in amyloid-related processes, other parameters and mechanisms are likely responsible for the variability in neuronal loss and tau pathology. These findings align with previous research indicating that the *APOE* genotype has a differential impact on A*β* between sexes ^33–35^. Understanding these sex-specific differences and the dominant influence of *APOE* status is essential to develop personalized therapeutic strategies that consider individual variability in disease progression.

### Limitation

There are a several limitations to our work. When generating our virtual sample, we assume that the parameters are independently distributed. This might not be the case in reality, as there might be correlation in biological features corresponding to parameters in a real population. Furthermore, we assume that the relative spread in the distribution of each parameter is the same. This might not be realistic, as some biological features are more homogeneous than others within a population. Another limitation is that outcome weighting was used during model calibration, and model performance may differ in the absence of weighting.

In addition, external validation of the complete model outputs was constrained by the availability of datasets containing all relevant biomarkers. While the UK Biobank cohort enabled validation of neuronal density (N) predictions at a large population scale, comparable A*β* and tau measurements were not available. Independent cohorts with A*β* and tau measurements will therefore be needed to further validate these model predictions.

A further limitation is that chronological age was used as the temporal variable to represent biological changes throughout the lifespan, and the current model does not explicitly incorporate an individualized disease progression time axis. However, this does not represent a fundamental limitation for the claims made in the present study, as the model was evaluated primarily at the population level. Although appropriate for modeling normal brain aging trajectories, chronological age does not fully capture individual differences in the timing and progression of pathological processes. The model nevertheless provides a framework from which individual biological trajectories can be extracted. Future work will focus on translating these trajectories into clinically meaningful measures of disease progression, such as individualized rates of progression and time-to-event estimates for clinically relevant outcomes, including dementia onset. Approaches such as the Disease Progression Score have been proposed to account for individual differences in disease onset and progression ^36^. Future developments incorporating personalized measures of disease progression might enable the model to better represent individual trajectories.

Finally, although sex and gender are not limited to binary categories, our current analysis is restricted to women and men because the available datasets provide only these two sex categories. Some model parameters are also defined using sex-specific values, while others are shared across groups. Future model development could incorporate a broader representation of sex-related biological variability as appropriate data and biological parameterization become available.

### Conclusion

This study presents the characterization, calibration, and validation of a mechanistic mathematical model of healthy brain aging, demonstrating its ability to reproduce key biological outcomes, amyloid beta (A*β*), tau (*τ*) and neuron density (N) across independent cohorts. The model showed strong agreement with observed neuronal density in the independent UK Biobank cohort, while calibration improved the prediction accuracy for A*β* and tau. Variance decomposition highlighted the contribution of *APOE* status to variability in A*β* and revealed sex-dependent differences in its influence on amyloid accumulation. The relatively weak correlations observed for some biomarkers suggest that brain aging trajectories arise from complex nonlinear interactions among multiple biological processes rather than a single dominant mechanism. Overall, these findings support the use of integrative computational models to study healthy brain aging and the potential emergence of AD-related pathological changes. By combining biological mechanisms with population-level variability, this framework provides a valuable tool for investigating how genetic and demographic factors influence age-related changes associated with Alzheimer’s disease risk, while providing future studies extending toward disease-related applications.

## Acknowledgements

Data used in the preparation of this article were obtained from the Alzheimer’s Disease Neuroimaging Initiative (ADNI) database (https://adni.loni.usc.edu).

Data collection and sharing for this project was funded by the Alzheimer’s Disease Neuroimaging Initiative (ADNI) (National Institutes of Health Grant U01 AG024904) and DOD ADNI (Department of Defense award number W81XWH-12-2-0012). ADNI is funded by the National Institute on Aging, the National Institute of Biomedical Imaging and Bioengineering, and through generous contributions from the following: AbbVie, Alzheimer’s Association; Alzheimer’s Drug Discovery Foundation; Araclon Biotech; BioClinica, Inc.; Biogen; Bristol-Myers Squibb Company; CereSpir, Inc.; Cogstate; Eisai Inc.; Elan Pharmaceuticals, Inc.; Eli Lilly and Company; EuroImmun; F. Hoffmann-La Roche Ltd and its affiliated company Genentech, Inc.; Fujirebio; GE Healthcare; IXICO Ltd.; Janssen Alzheimer Immunotherapy Research & Development, LLC.; Johnson & Johnson Pharmaceutical Research & Development LLC.; Lumosity; Lundbeck; Merck & Co., Inc.; Meso Scale Diagnostics, LLC.; NeuroRx Research; Neurotrack Technologies; Novartis Pharmaceuticals Corporation; Pfizer Inc.; Piramal Imaging; Servier; Takeda Pharmaceutical Company; and Transition Therapeutics. The Canadian Institutes of Health Research is providing funds to support ADNI clinical sites in Canada. Private sector contributions are facilitated by the Foundation for the National Institutes of Health (https://www.fnih.org). The grantee organization is the Northern California Institute for Research and Education, and the study is coordinated by the Alzheimer’s Therapeutic Research Institute at the University of Southern California. The ADNI data are disseminated by the Laboratory for Neuro Imaging at the University of Southern California.

This research also used data from the UK Biobank Resource (https://pubmed.ncbi.nlm.nih.gov/25826379/) under application number 85063. UK Biobank is an open-access biomedical research resource that provides researchers with approved access to health, genetic, and imaging data for scientific research.

## Ethical considerations

This work involved secondary analysis of de-identified human data from the Alzheimer’s Disease Neuroimaging Initiative (ADNI) and the UK Biobank (UKBB) under application number 85063. All original studies obtained informed consent from participants and received approval from their institutional review boards. No additional human or animal subjects were enrolled in this research.

## Consent to participate

Informed consent was obtained from all participants in the original ADNI and UK Biobank studies. The present study involved only secondary analysis of de-identified data obtained from these established cohorts, and no additional participant recruitment or data collection was performed.

## Data availability statement

Data supporting the findings of this study are openly available from UKBB (https://www.ukbiobank.ac.uk/) and ADNI (https://adni.loni.usc.edu/) upon formal application and approval.

## A Appendix: Equations for Microglia, Macrophages, Cytokines, and Chemokines

This appendix presents equations for amyloid beta, NFTs, dead neurons, microglia, macrophages, cytokines, and chemokines that were taken from previous work ^15^. The equations have been summarized for clarity.

### A.1 Equations for amyloid beta

#### A.1.1 Equation for intracellular amyloid beta monomers

The equation for monomeric A*β* is:

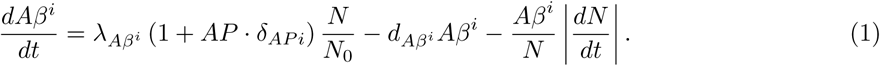

This equation describes the creation, degradation, and externalization of intracellular A*β*.

#### A.1.2 Equation for extracellular amyloid beta monomers

The equation for extracellular A*β* monomers is:

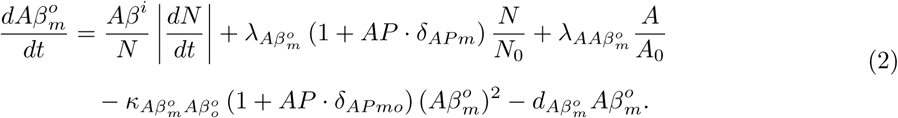

This equation represents the transition from intracellular to extracellular amyloid, creation of extracellular A*β* monomers, production by astrocytes, aggregation into oligomers, and degradation. The squared term in the aggregation process is based on the findings of ^37^.

#### A.1.3 Equation for extracellular amyloid beta oligomers

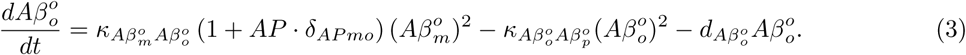

This equation describes the aggregation of monomers into oligomers, oligomers into plaques, and oligomers degradation.

#### A.1.4 Equation for extracellular amyloid beta plaques

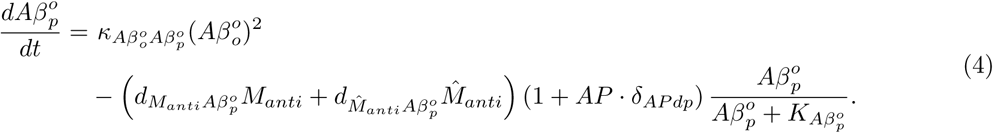

This equation represents oligomer-to-plaque conversion and plaque degradation by anti-inflammatory microglia and macrophages. Activated macrophages and microglia can eliminate A*β* plaques ^38–42^.

### A.2 Equation for glycogen synthase kinase 3 (GSK-3)

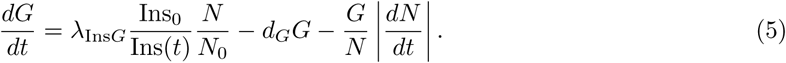

This equation describes the creation and degradation of GSK-3, influenced by insulin concentration. GSK-3*β* is more dysregulated in AD ^43^, and its activity is modulated by insulin concentration ^44–46^.

### A.3 Equation for phosphorylated/hyperphosphorylated tau proteins

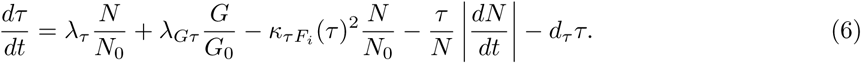

This equation describes the production, aggregation, release, and degradation of hyperphosphorylated tau proteins. Tau proteins in AD patients are three to four times more phosphorylated than in age-matched individuals without cognitive difficulties ^47^.

### A.4 Equations for NFTs

#### A.4.1 Intracellular NFTs

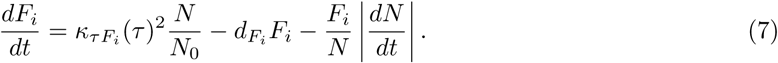

This equation represents the formation, degradation, and release of intracellular neurofibrillary tangles (NFT).

#### A.4.2 Extracellular NFTs

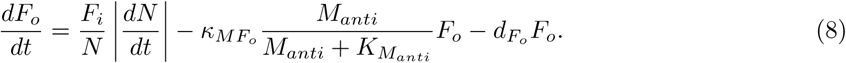

This equation describes the release, degradation by microglia, and other degradation processes of extracellular NFTs.

### A.5 Equation for the density of neurons

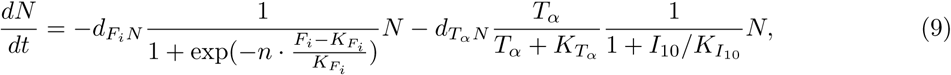

This equation represents intracellular NFTs and proinflammatory cytokines as the cause of neuronal death, with anti-inflammatory cytokines as the inhibitor.

### A.6 Equation for the density of activated astrocytes

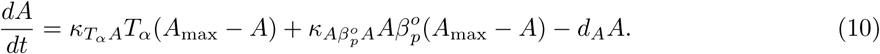

This equation describes the activation of astrocytes by TNF-*α* and A*β* plaques, and their deactivation and death. Astrocytes are activated by TNF-*α* and A*β* plaques ^48–50^.

### A.7 Equations for microglia

#### Resting microglia

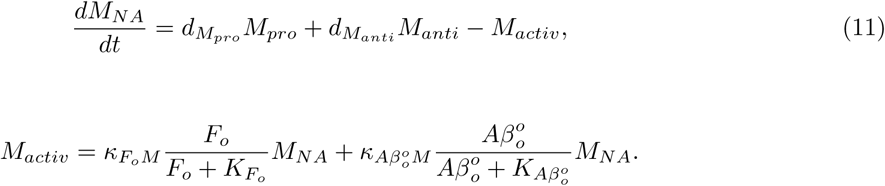

These equations describe the dynamics of resting microglia, including activation by extracellular NFTs and A*β* oligomers ^41;51–53^.

#### Activated microglia

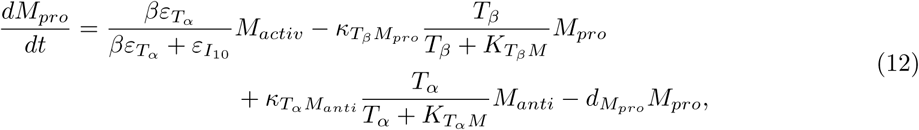

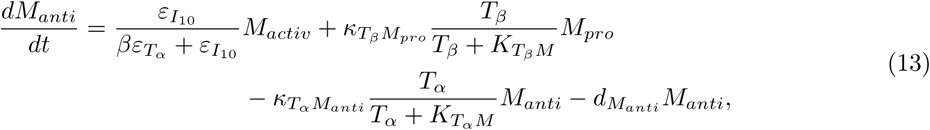

These equations describe the dynamics of activated pro-inflammatory and anti-inflammatory microglia, including polarization and conversion between states ^41;42;54–56^.

### A.8 Equations for activated macrophages

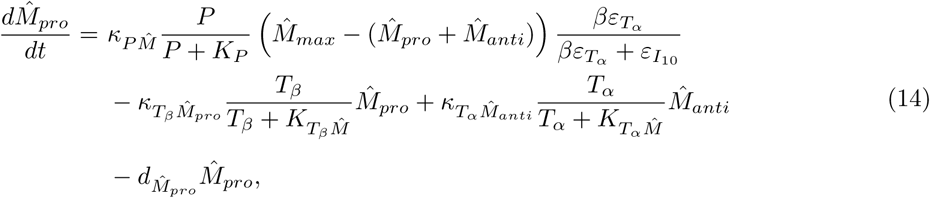

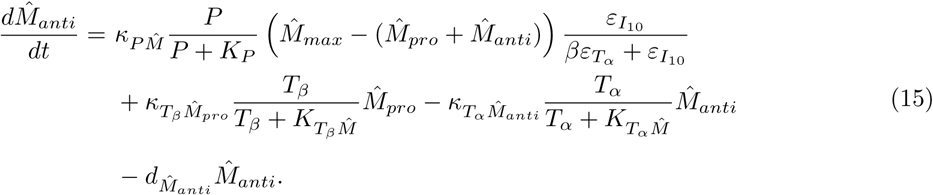

These equations describe the dynamics of pro-inflammatory and anti-inflammatory activated macrophages, including polarization, conversion between states, and degradation ^40–42;54–59^.

### A.9 Equations for cytokines and chemokines

Our model also describes the evolution of relevant cytokines and chemokines, including TGF-*β*, IL-10, TNF-*α*, and MCP-1.

#### A.9.1 Transforming growth factor beta

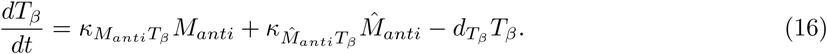

This equation describes the production and degradation of TGF-*β* by anti-inflammatory microglia and macrophages ^41;55;60^.

#### A.9.2 Interleukin 10

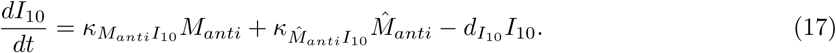

This equation describes the production and degradation of IL-10 by anti-inflammatory macrophages and microglia ^41;55;60^.

#### A.9.3 Tumour necrosis factor-alpha

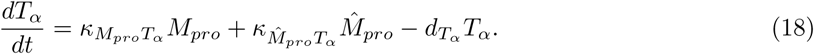

This equation describes the production and degradation of TNF-*α* by pro-inflammatory microglia and macrophages ^48;49;60^.

#### A.9.4 Monocyte chemoattractant protein-1

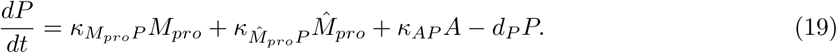

This equation describes the production and degradation of MCP-1 by pro-inflammatory macrophages, microglia, and activated astrocytes ^55;58;61^.

## B Appendix: Sources of Model Optimized Parameters

This appendix lists all the optimized parameters used in the equations for amyloid-beta, NFTs, dead neurons, microglia, macrophages, cytokines, and chemokines, sourced from the literature.

| Parameter | Signification | Source |
| --- | --- | --- |
| **Amyloid-beta ($A\beta$)** |  |  |
| $\lambda_{AB_m^o}$ | $A\beta$ monomer creation outside neuron | 31;62 |
| $\kappa_{AB_m^o AB_o^o}$ | Monomer to oligomer conversion rate | 37 |
| $\delta_{AP_{mo}}$ | APOE4 effect on monomer-oligomer conversion | 63 |
| $\kappa_{AB_{oo} \rightarrow AB_{po}}$ | Oligomer to plaque conversion rate | 64;65 |
| $d_{AB_o^o}$ | Oligomer degradation rate | 37 |
| $d_{M_{antiAB_p^o}}$ | Plaque degradation by anti-inflammatory microglia | 66 |
| $\delta_{AP_{dp}}$ | APOE4 effect on plaque degradation | 67 |
| $K_{AB_p^o}$ | Michaelis-Menten constant for plaque degradation | 68 |
| **GSK-3, tau protein and NFTs** |  |  |
| $Ins_0$ | Normal insulin concentration in brain at 30 years old | 15 |
| $d_G$ | Degradation rate of GSK-3 | 69 |
| $G_0$ (men) | Normal GSK-3 concentration, based on brain density | 70 |
| $G_0$ (women) | Normal GSK-3 concentration, based on brain density | 71 |
| $\lambda_{InsG}$ (men) | Creation rate of GSK-3 for men induced by insulin | 69;70 |
| $\lambda_{InsG}$ (women) | Creation rate of GSK-3 for women induced by insulin | 69;71 |
| $\lambda_{G\tau}$ | Tau phosphorylation by GSK-3 | 72;73 |
| $\kappa_{\tau F_i}$ | Conversion rate of tau to intracellular NFTs | 74–76 |
| $d_{\tau}$ | Tau degradation rate, based on half-life | 77 |
| $d_{F_i}$ | Intracellular NFT degradation | 31;78 |
| **Neurons** |  |  |
| $d_{F_i N}$ | Max neuron death rate from $F_i$ | 79 |
| $K_{F_i}$ | Half-max concentration of $F_i$ for neuron death | 80 |
| $d_{T\alpha N}$ | Max neuron death rate from TNF-α | 81 |
| **Microglia** |  |  |
| $\kappa_{T_b M_{pro}}$ | Concentration of Aβ oligomers extracellular at which microglia activation is half maximal | 82 |
| $K_{T_B M}$ | Half saturation constant of TGF-β for $M_{anti}$ to $M_{pro}$ conversion | 83–85 |
| $\kappa_{T_{\alpha} M_{anti}}$ | Max rate of conversion $M_{anti}$ to $M_{pro}$ under signaling of TNF-α | 41;82 |
| $K_{T_{\alpha} M}$ | Half-saturation constant of TNF-α for $M_{anti}$ to $M_{pro}$ conversion | 86 |
| **Macrophages** |  |  |
| $\kappa_{T_{\alpha} \hat{M}_{anti}}$ | Maximum rate of conversion from anti-inflammatory to pro-inflammatory macrophages ($\hat{M}_{anti} \rightarrow \hat{M}_{pro}$) under TNF-α signaling | 41;87 |
| **Cytokines and Chimiokines** |  |  |
| $\kappa_{M_{anti} T_{\beta}}$ | TGF-β production rate by anti-inflammatory microglia | 88;89 |
| $\kappa_{\hat{M}_{anti} T_{\beta}}$ | TGF-β production rate by anti-inflammatory macrophages | 88;89 |
| $d_{T_{\beta}}$ | TGF-β degradation rate | 90 |
| $d_{T_\alpha}$ | TNF-α degradation rate | 91;92 |
| $\kappa_{M_{\text{pro}}T_\alpha}$ | TNF-α production rate by pro-inflammatory microglia | 49;60;89 |
| $\kappa_{\hat{M}_{\text{pro}}T_\alpha}$ | TNF-α production rate of pro-inflammatory macrophages | 49;60;89 |

